# Impact of National and Regional Lockdowns on Growth of COVID-19 Cases in COVID-Hotspot City of Pune in Western India: A Real-World Data Analysis

**DOI:** 10.1101/2021.05.05.21254694

**Authors:** Vidya Mave, Arsh Shaikh, Joy Merwin Monteiro, Prasad Bogam, Bhalchandra S Pujari, Nikhil Gupte

## Abstract

**Background:** Real-world data assessing the impact of lockdowns on COVID-19 cases remain limited from resource-limited settings. We examined growth of incident confirmed COVID-19 cases before, during and after lockdowns in Pune, a city in western India with 3.1 million population that reported the largest COVID-19 burden at the peak of the pandemic.

**Methods:** Using anonymized individual-level data captured by Pune’s public health surveillance program between February 1^st^ and September 15^th^ 2020, we assessed weekly incident COVID-19 cases, infection rates, and epidemic curves by lockdown status (overall and by sex, age, and population density) and modelled the natural epidemic using the 9-compartmental model INDSCI-SIM. Effect of lockdown on incident cases was assessed using multilevel Poisson regression. We used geospatial mapping to characterize regional spread.

**Findings:** Of 241,629 persons tested for SARS-CoV-2, the COVID-19 disease rate was 267.0 (95% CI 265.3 – 268.8) per 1000 persons. Epidemic curves and geospatial mapping showed delayed peak of the cases by approximately 8 weeks during the lockdowns as compared to modelled natural epidemic. Compared to a subsequent unlocking period, incident COVID-19 cases 43% lower (IRR 0.57, 95% CI 0.53 – 0.62) during India’s nationwide lockdown and 22% (IRR 0.78, 95% CI 0.73 – 0.84) during Pune’s regional lockdown and was uniform across age groups and population densities.

**Conclusion:** Lockdowns slowed the growth of COVID-19 cases in population dense, urban region in India. Additional analysis from rural and semi-rural regions of India and other resource-limited settings are needed.

## Introduction

Worldwide, the trajectory of COVID-19 cases caused by SARS-CoV-2 has continued to rise since first detected in December 2019 in Wuhan, China.[1-3] As of March 1st, 2021, global cases exceeded 116 million with USA reporting the largest caseload and India – the world’s second most populous country – recording the second largest cumulative caseload at 11.2 million.[1] With mounting morbidity and mortality in resource-rich and resource-limited settings alike[1, 3-7], scientists across the globe have developed vaccines and investigating therapeutics and vaccines against COVID-19.[8-12] However, alternative non-pharmacological strategies are critical to limit COVID-19 transmission.

Modelling exercises from India and elsewhere support the use of several such interventions to contain the spread of the COVID-19 pandemic, including complete lockdowns, curfews, regional containment strategies, social distancing, and the strict use of barrier protections (i.e., gowning, use of masks, handwashing, etc.).[13-18] Among these, complete lockdowns are most recommended. Several European countries have adopted this strategy with relatively high success rates and more recent re-implementation. However, few assessments report the impact of lockdown in resource-limited settings, including India, which comprises large variations in urban and rural population density^19-22^

Early in the COVID-19 pandemic, India instituted a nationwide lockdown for an extended 68-day period, which was followed by staggered phases of relaxation.[19] Notably, a rise in new confirmed cases prompted a second, regional lockdown in Pune city, a tier 2 city in western India. Using public COVID-19 surveillance data collected between February 1^st^ and September 15^th^, 2020, we aimed to assess the real-world impact of lockdown on incident COVID-19 cases in Pune city municipality and its subregions of variable population density. Further, we aimed to characterize the geospatial spread of the cumulative COVID-19 burden.

## Methods

### COVID-19 surveillance program in Pune, India

Pune city − located in western India around 150 kilometers east of Mumbai (**Figure 1A**) − launched a COVID-19 surveillance program during the early stages of the pandemic (January 2020). Pune Municipal Corporation (PMC) collaborated with multiple public and private health facilities to establish SARS-CoV-2 diagnostics, quarantine facilities for asymptomatic persons, and hospital/critical care beds for moderate to severely ill patients diagnosed with COVID-19. In addition, community-based workers were mobilized to conduct contact tracing activities. A publicly accessible dashboard was established to report the cumulative COVID-19 caseload in the PMC’s 41 *Prabhags* (also known as electoral wards). The number tested and individual-level data, such as age, sex, residential address, COVID-19 test results, and COVID-19 outcomes, were centrally compiled on a regular (almost daily) basis.[20]

**Figure 1.**
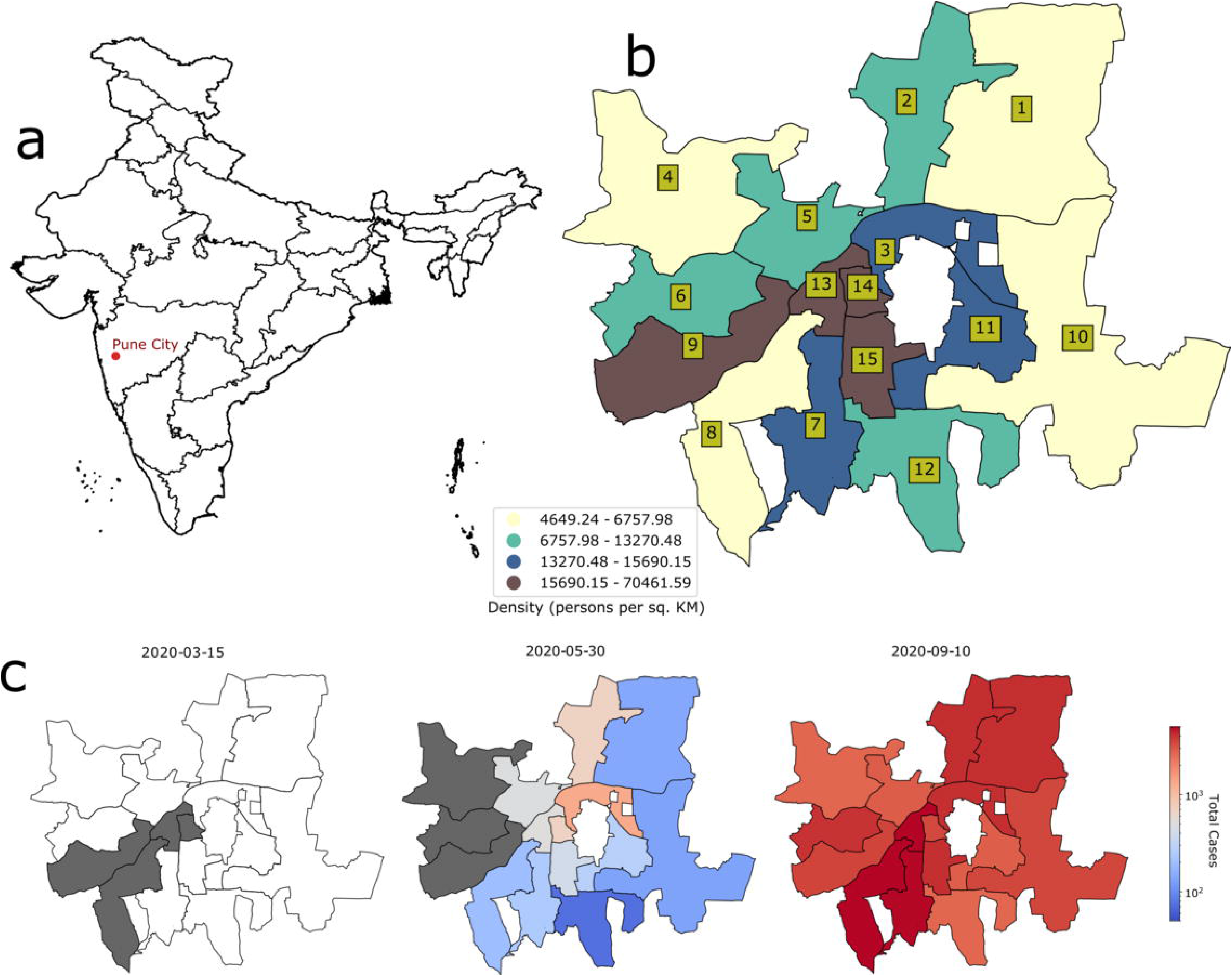
**A** Location of Pune City, India. **1B** Geographic boundaries of ward offices located within Pune Municipal Corporation (PMC). Fill color indicates the quartile of population density (persons per square kilometer). Numbers inside the olive boxes indicate the official ward office number (see **Supplemental Table S1** for the name corresponding to each ward office number). **1C** the number of COVID-19 cases in each PMC ward office at beginning of the pandemic **(left panel)**, at the end of the nationwide lockdown **(middle panel)**, and at the end of the study period **(right panel)**. The date is located at the top of each panel. Dark gray indicates <50 cases, white indicates no cases, and the transition between blues and reds seen in the middle panel denotes approximately 600 cases.

### Nationwide and Pune regional COVID-19 pandemic management

India’s initial response to the pandemic comprised travel advisories on international travel and suspension of visas from mid-January through mid-March. During this period, COVID-19 testing was administered to travelers who were returning from China and other foreign countries and had fever, cough or other viral respiratory symptoms.^19^ Those testing positive were hospitalized for quarantine, and their close contacts were traced and underwent COVID-19 testing. The first nationwide lockdown was implemented from March 25^th^ to April 14th, 2020 (Lockdown 1). Nearly all services and factories were suspended with reports of arrests for lockdown violations. During this time, Pune city expanded COVID-19 testing capacity, making testing available to persons with viral symptoms or within 14 days of COVID-19 exposure. The nationwide lockdown was extended from April 15^th^ to May 3^rd^ (Lockdown 2). Agricultural activities and essential services were allowed to function on April 20th, and Pune city areas were classified into red, orange, and green zones based on infection clusters. Red zones were defined by the central government based on case counts, doubling rate, and testing/surveillance findings. Green zones were areas where cases had never been reported or where there had been no reported cases in 21 days. Last, orange zones were areas deemed neither a red zone nor a green zone by the central government. Notably, interstate transport was allowed for stranded individuals, and during the month of May alone, approximately one million migrants traveled via roads or trains to their home states, mostly from Maharashtra state. The lockdown was extended again from May 4^th^ to May 17^th^ (Lockdown 3), but with more relaxations in green zones where lower infection rates were reported. The final extension spanned May 18^th^ to May 31^st^ (Lockdown 4). States were given more authority to demarcate infection zones, and red zones were further divided into containment zones, which maintained stricter enforcement of lockdown norms than other zones.

The unlocking (resumption) of economic activities began in June 2020. During the first phase (Unlock 1, June 1^st^ to June 30th), interstate travel was allowed with few state-specific restrictions while containment zones continued to follow lockdown norms. Phased unlocking continued in July (Unlock 2) when the authority to impose lockdowns was further decentralized to local governments. Pune city and the adjoining areas implemented a regional lockdown from July 14^th^ to July 23^rd^ in response to a sharp rise in COVID-19 cases. City and state authorities enforced a strict lockdown during the first week – a complete shutdown of all essential services, except emergency healthcare. This resulted in minimal movement in Pune’s public spaces. Slight relaxations in the supply of essential goods and services followed during the second week and Unlock 2 resumed in Pune on July 24th. August 1^st^ to August 31^st^ (Unlock 3) witnessed further relaxations in interstate travel and an end to nationwide curfews. Pune shopping malls and market complexes could remain open until evening, and cab services could operate with a restricted passenger load. However, lockdown restrictions continued in containment zones. During September (Unlock 4), gatherings of up to 50 persons were permitted while containment zones continued to follow lockdown norms. Early in September, Pune experienced a sharp rise in COVID-19 cases and became a top national COVID-19 hotspot.

### Data curation

The area within PMC limits is divided into 15 administrative units, called ward offices (**Figure 1B**), which are further divided into 41 electoral wards with similar populations, called *prabhags*. Individual-level data were included for the time period spanning February 1^st^ to September 15^th^, 2020. According to daily press reports released by PMC, a total of 542946 samples were collected for COVID-19 testing during the study period, and of these, 313373 records were available. These data were curated to remove records with missing data. The remaining records were assigned to a *prabhag* using a machine learning based geocoder that was developed in house. The geocoding methodology is described in the supplementary material 1. Records with a confidence score below 0.5 out of 1.0 (provided by the ML geocoder) and records for persons residing outside PMC limits were removed. The final dataset used for this analysis comprises 241629 records. Ethics Committee of Indian Institute of Science Education and Research, Pune, India approved the analysis of COVID-19 programmatic data.

### Healthcare preparedness data

Publicly available data on key health system metrics were abstracted from newspapers (as available) immediately following lockdown and updated just prior to the end of the analysis period. Metrics included number of flu clinics, number of available SARS CoV-2 tests, number of hospital beds, and number of critical care beds.

### Statistical analysis and mathematical modelling

The primary endpoint was weekly change in incident COVID-19 cases. The secondary endpoint was weekly infection rate; infection rate was calculated as the number of positive SARS-CoV-2 results divided by the total number of tests per 1000 population. Other endpoints included risk of COVID-19. Primary and secondary endpoints were assessed pre-lockdown, during lockdown and post-lockdown in the overall dataset and by population characteristics, namely sex, age group, and ward office-specific subcategories (population density and proportion residing in slum areas). Population density was calculated as number of people per 1 square kilometer and has been reported for all 15 PMC ward offices. For this analysis, population density was binarized as high (above the 3^rd^ quartile of PMC ward office density, n=6) or low-average (below the 3^rd^ quartile of PMC ward office density, n=9) (**Figure 1B)**. Since differences in infection rates existed among ward offices, the effect of lockdown on the primary endpoint was assessed using a multilevel Poisson regression model with random effects for ward office and test week. Change in the weekly infection rate over the study period was estimated using quasi-Poisson regression analysis. Logistic regression was used to assess the effect of risk factors on mortality. Epidemic curves for trends of incident cases over time were plotted using nonparametric locally weighted regression for the overall population and by sex, age group, and ward-specific subcategories.

We modelled the trajectory of the natural epidemic to estimate the delay of the peak of the pandemic. For this, we used a 9-compartmental model INDSCI-SIM that enables robust predictions taking into account the effects of various non-pharmaceutical measures[21, 22]. In order to predict the natural trajectory of the epidemic in absence of any lockdown, we set all the parameters to their default values except for the effective reproduction number *R*_*0*._ [21] There are wide range of estimates for the value of *R*_*0*_; for example Hilton and Keeling estimated *R*_*0*_ between 2 and 3[23] while India specific study by Sinha found out the value to be around 1.8. In order to avoid overestimation of total cases, we also considered *R*_*0*_ =1.8 [24]. Although there is no unique way to estimate actual number of cases, we assume infection on the first day (taken to be 1st April 2020) of the simulation to be three times reported cases. We note here that the choice of *R*_*0*_ and initial values may affect the final outcome, but our choices are conservative and more accurate estimation may make the results worse that reported here. We assessed the geospatial spread of COVID-19 cases over time using the Python library geopandas (version 0.7.0). Data were analyzed in Stata Version 14.2.

Ethics Committee of Indian Institute of Science Education and Research, Pune, India approved the analysis of COVID-19 programmatic data.

## Results

### Population characteristics and risk for COVID-19 and mortality

From February 1^st^ to September 15^th^, 2020, of 241,629 SARS CoV-2 tests performed in all 15 PMC ward offices, 64,526 (26%) were positive, contributing to an overall rate of COVID-19 disease of 267.0 (95% CI 265.3 – 268.8) per 1000 persons. Among those diagnosed with COVID-19 disease, the median age was 36 (interquartile range [IQR] 25 – 50) years, 36,180 (56%) were male, and 9414 (15%) were children <18 years. Compared to persons ages <5 years, risk of contracting COVID-19 was higher among ages 5 – 18 years (incidence risk ratio [IRR] 1.11, 95% CI 1.06 – 1.17), 18 – 35 years (IRR 1.13, 95% CI 1.08 – 1.19), 35-50 years (IRR 1.2, 95% CI 1.23 – 1.35) and >50 years (IRR 1.50, 95% CI 1.43 – 1.57) (**Table 1**).

**Table 1.**
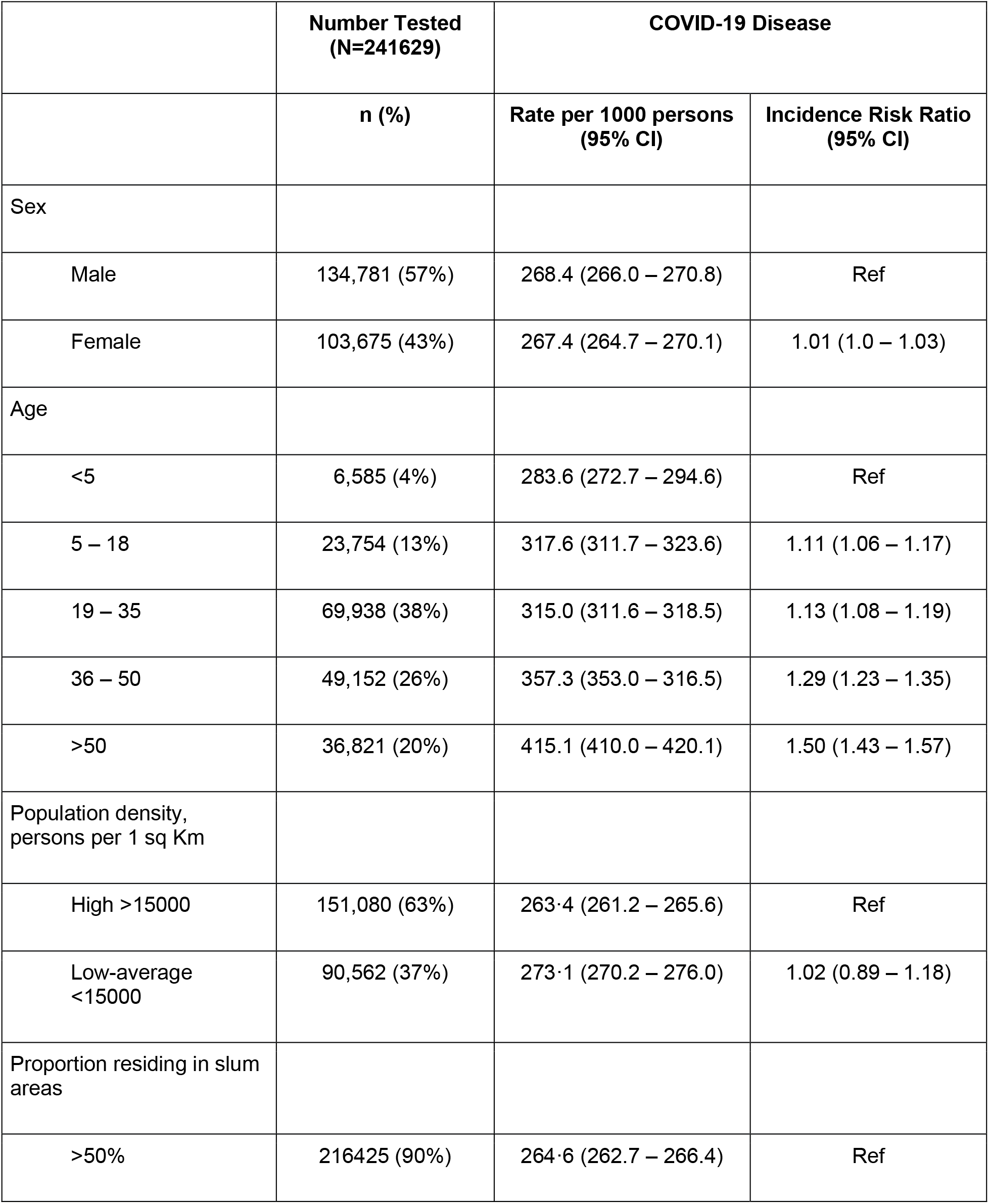
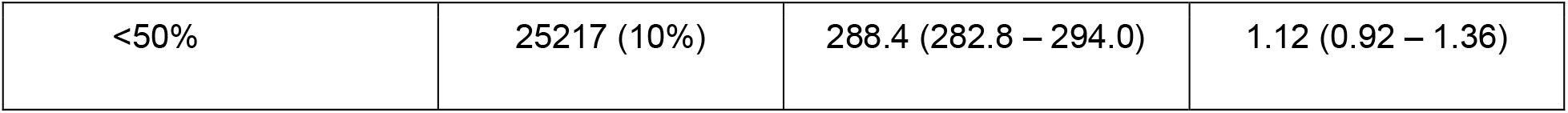
Estimated rate and incidence risk ratio of COVID-19 disease during the study period by population characteristics in Pune Municipal Corporation, India.

### Lockdowns and incident COVID-19 cases

**Figure 2A-B** illustrates the overall trajectory of new COVID-19 cases over the study period by lockdown status. In our model of the natural epidemic model, the epidemic would have reached the peak in mid-July as against to actual peak in mid-September in the absence of national lockdown. The approximate delay of the growth of the cases was 8 weeks (Figure **2B**). New COVID-19 cases maintained a steady rise before and during the nationwide lockdown with a 5% (1% – 8%) weekly increase in new infections during the lockdown (**Table 2**). This trajectory leveled off during the regional lockdown with a 7% (−17% – 4%) weekly decrease in new infections (**Table 2**). Subsequent unlock periods (unlock 2 – 4) witnessed a sharp rise in incident COVID-19 cases with the largest weekly increase in new infections of 9% (7% – 10%) (**Table 2**). Incident cases peaked around the first week of September when Pune reported India’s largest burden of active COVID-19 disease (**Figure 2AB, p < 0**.**001**)(**Table 2**).

**Table 2.** Percent change (95% CI) in weekly incident COVID-19 infections by lockdown phase in the overall dataset and by population characteristics^a^.

|  | **Pre-Lockdown** | **Nationwide Lockdown** | **Unlock 1 – 2** | **Regional Lockdown** | **Unlock 2** | **Unlock 3 – 4** |
| --- | --- | --- | --- | --- | --- | --- |
| Overall | -1% (-18% – 19%) | 5% (1% – 8%) | 0.4% (0.2% - 3%) | -7% (-17% – 4%) | -2% (-11% – 8%) | 9% (7% – 10%) |
| Sex |  |  |  |  |  |  |
| Male | -3% (-17% – 14%) | 3% (-1% - 6%) | 0.4% (-2% - 3%) | -6% (-17% - 6%) | -3% (-17% - 15%) | 10% (8% - 11%) |
| Female | -4% (-27% – 27%) | 4% (-0.3% - 7%) | 0.3% (-3% - 3%) | -8% (-17% - 3%) | 1% (-12% - 15%) | 8% (6% - 10%) |
| Age |  |  |  |  |  |  |
| <18 years | 38% (-7% - >95%) | 2% (-1% – 5%) | -21% (-23% – -19%) | -7% (-17% - 4%) | -8% (-21% - 7%) | 10% (8% - 12%) |
| ≥18 years | -2% (-13% - 10%) | -1% (-2% – 1%) | 2% (1% – 3%) | -6% (-10% – -1%) | -2% (-8% – 5%) | 9% (8% - 10%) |
| Population density, persons per sq Km |  |  |  |  |  |  |
| High >15000 | -3% (-18% - 16%) | -2% (-7% – 2%) | 6% (2% - 9%) | -14% (-25% - -1%) | -8% (-22% – 8%) | 6% (3% - 9%) |
| Low <15000 | -3% (-38% - 52%) | 10% (5% - 14%) | -2% (-5% - 1%) | -3% (-22% - 20%) | 2% (-11% – 17%) | 10% (8% - 12%) |
| Proportion residing in slum areas |  |  |  |  |  |  |
| >50% | -14% (-48% - 43%) | 12% (-3% – 31%) | 2% (-6% - 2%) | -8% (-37% - 32%) | -13% (-37% - 20%) | 9% (6% - 12%) |
| <50% | 1% (-19% - 25%) | 4% (1% - 7%) | 1% (-2% - 4%) | -7% (-18% - 5%) | 1% (-15% - 20%) | 8% (7% - 10%) |

<sup>a</sup> Pre-Lockdown (Feb 1 to March 24, 2020); Nationwide Lockdown (March 25 to May 31, 2020); Unlock 1 – 2 (June 1 to July 13, 2020); Regional Lockdown (July 14 to July 23, 2020); Unlock 2 (July 24 to July 31, 2020); Unlock 3 –4 (September 1 to September 30, 2020).

**Figure 2.**
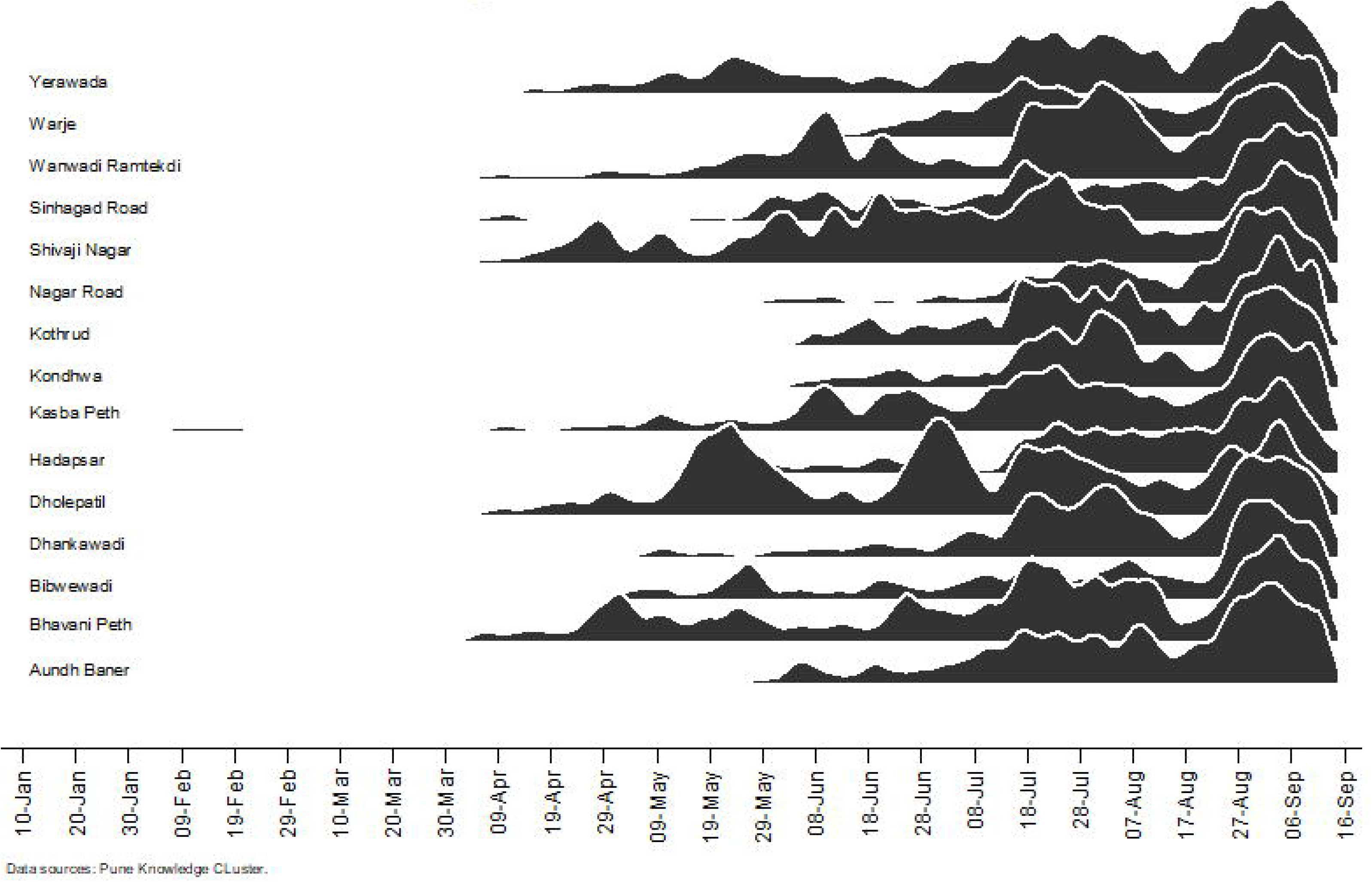
shows the trajectory of new COVID-19 cases within Pune Municipal Corporation (PMC) over time. Lockdown/unlock periods defined as: pre-lockdown (1st February to 24th March 2020); nationwide lockdown (25th March to 31st May 2020); unlock 1 – 2 (1st June-13th July 2020); Pune regional lockdown (14th July to 23rd July 2020); and unlock 2 – 4 (23rd July to September 15th, 2020). **Figure 2A.** Number of daily incident COVID-19 cases across Pune Municipal Corporation ward offices during the study period.

**Figure 2B.**
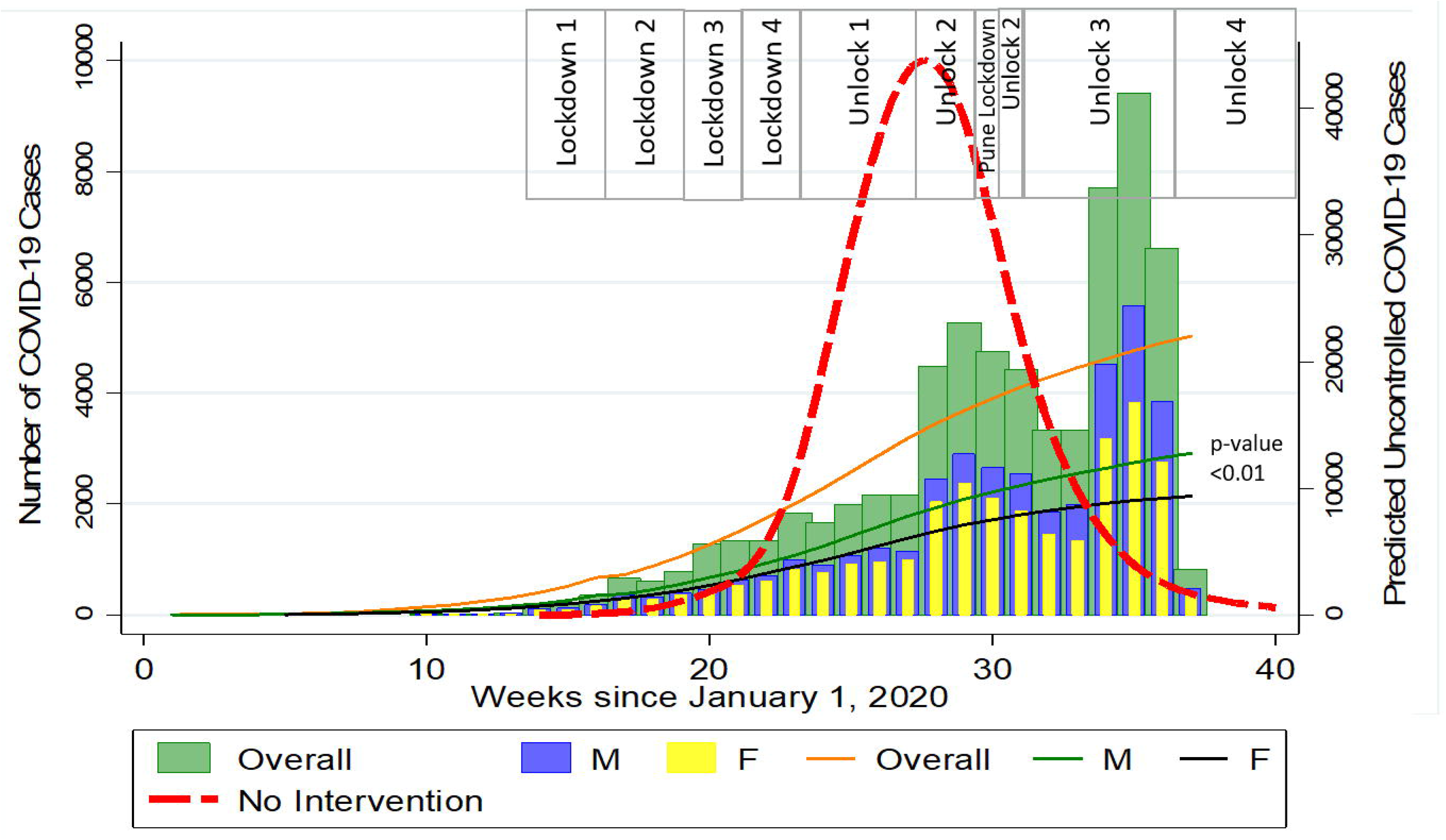
Weekly incident COVID-19 cases by natural epidemic, and lockdown status in the overall dataset and by sex. The red broken line represents projection modelling of the natural epidemic considering *R*_*0*_ =1.8; the orange line shows the weekly incident COVID-19 cases overall by lockdowns; the green and black line shows the incident COVID-19 cases among males and females, respectively.

Incident COVID-19 case trajectories by sex and age group illustrate a similar pattern (**Figure 2BC**)(**Table 2**). Ward offices with high population density (>15000 persons/square Km) had a significantly fewer new cases than wards with low-average population density (<15000 persons/square Km) (**Figure 2D, p < 0**.**001**). During the nationwide lockdown, high population density areas had the lowest new infection rates with a 2% (−7% – 2%) weekly decrease in new cases. Immediately following the Pune regional lockdown, areas with a majority living in slum areas had the lowest infection rate with a 13% (−37% – 20%) weekly decrease in new infections (**Table 2**).

**Figure 2C.**
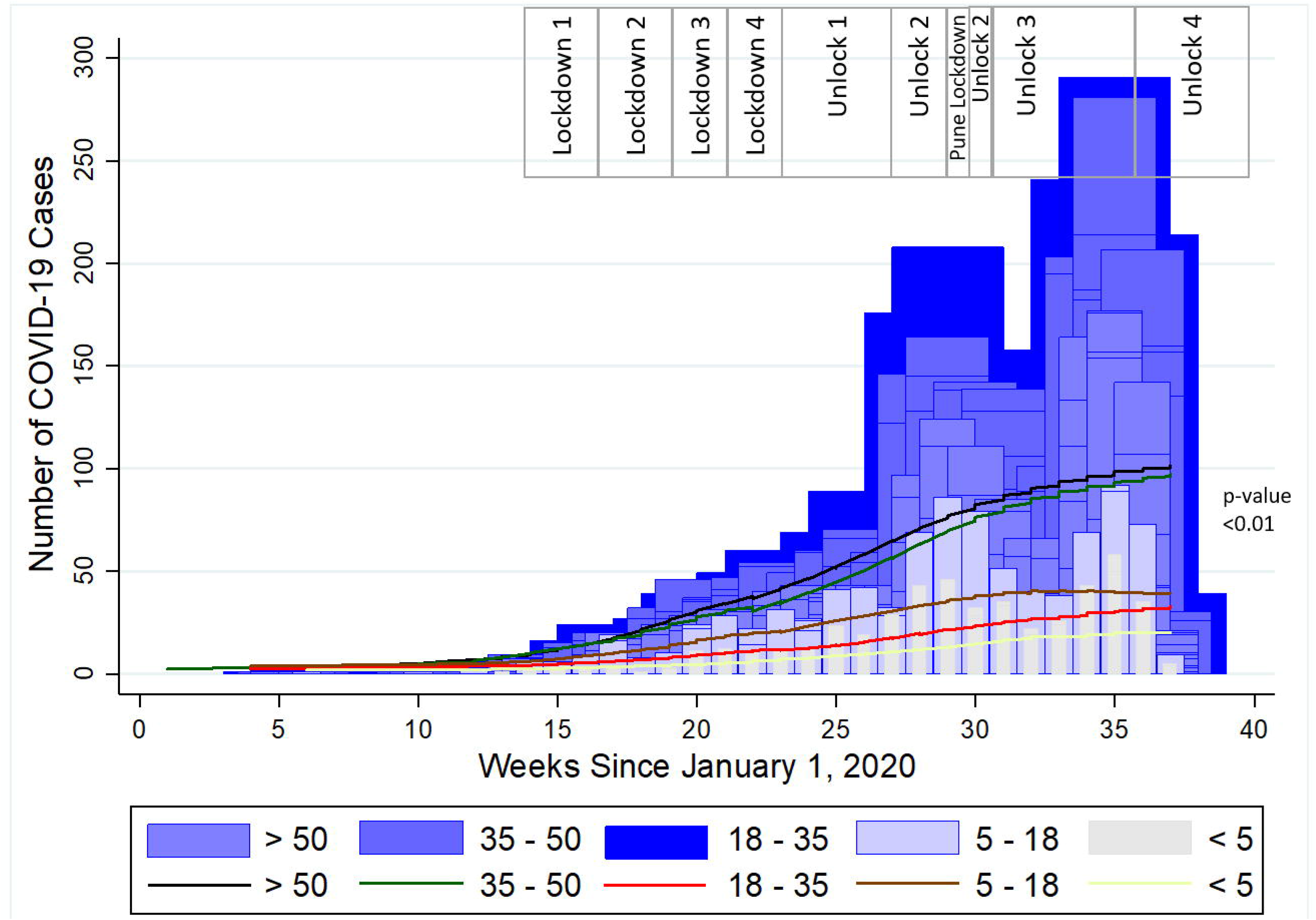
Weekly incidence of COVID-19 by age groups

**Figure 2 D.**
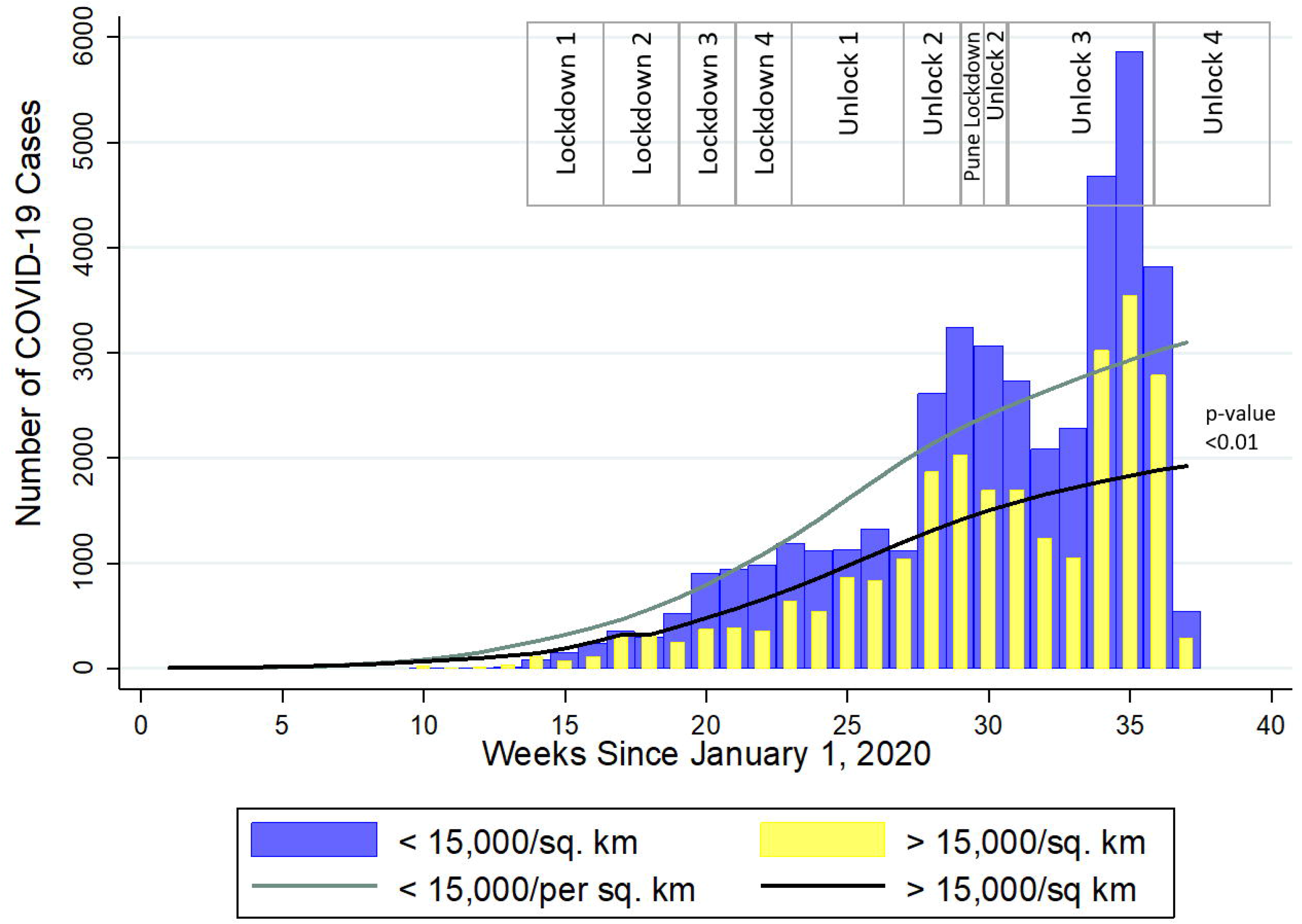
Weekly incident COVID-19 cases by population density.

In multilevel Poisson regression models (**Table 3**), incident COVID-19 cases were 43% lower during the nationwide lockdown (IRR 0.57, 95% CI 0.53 – 0.62) and 22% lower during the Pune regional lockdown (IRR 0.78, 95% CI 0.73 – 0.84) compared to the post-regional lockdown period (unlock 3 – 4). Similar trends were observed across population characteristics, including sex, age, population density, and proportion residing in slum areas.

**Table 3.**
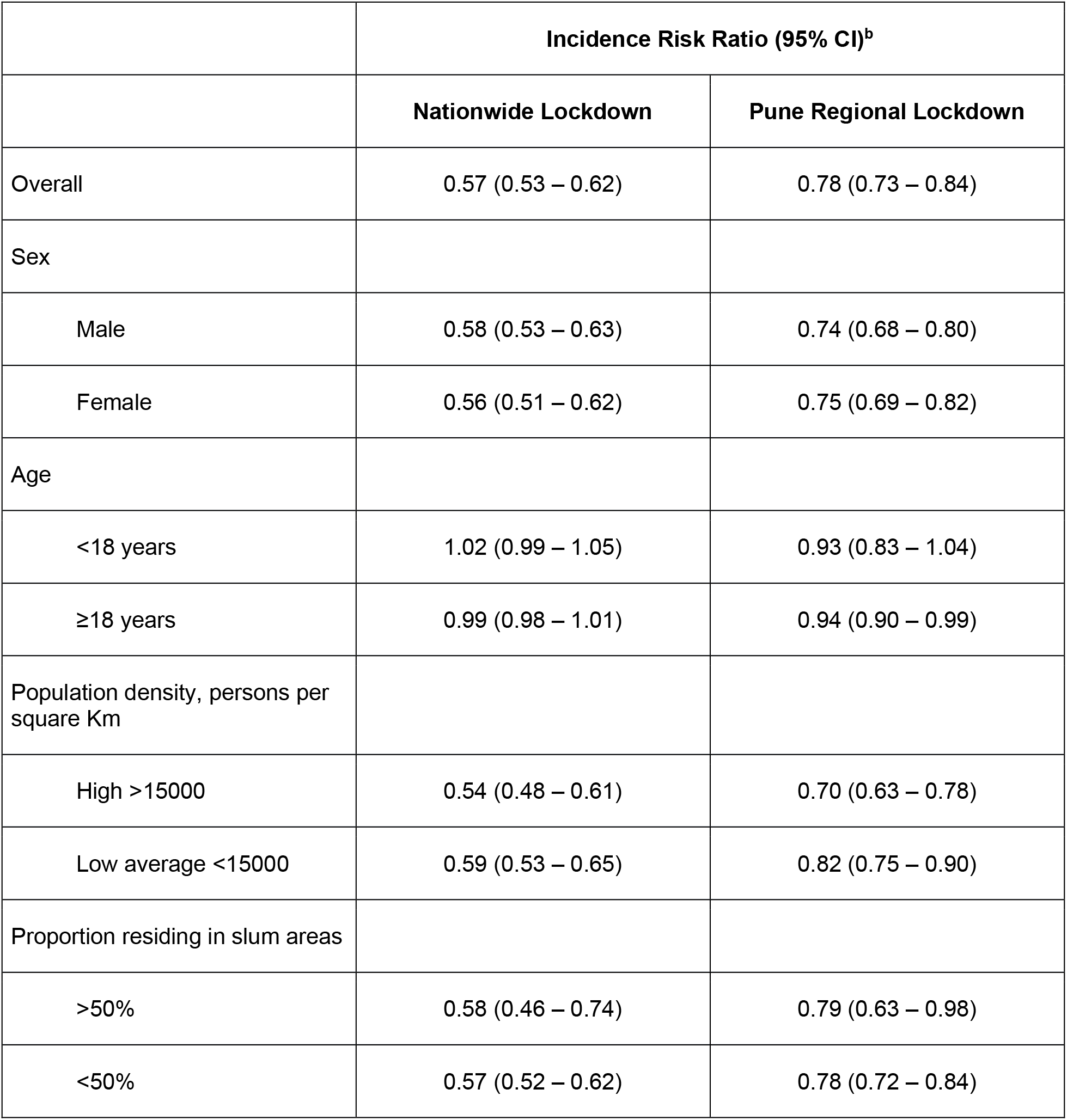
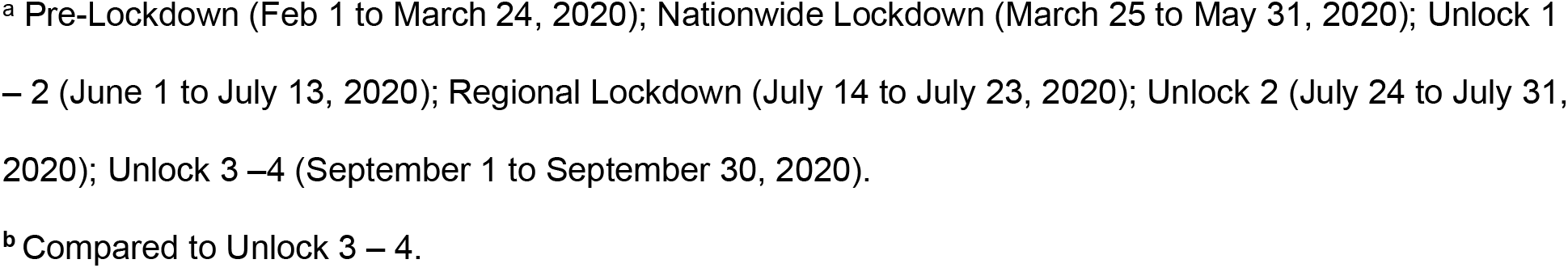
Estimated effect of nationwide and Pune regional lockdowns on weekly incident COVID-19 cases using Poisson regression in Pune India, overall and by population characteristics^a^.

|  | **Incidence Risk Ratio (95% CI)<sup>b</sup>** |  |
| --- | --- | --- |
|  | **Nationwide Lockdown** | **Pune Regional Lockdown** |
| Overall | 0.57 (0.53 – 0.62) | 0.78 (0.73 – 0.84) |
| Sex |  |  |
| Male | 0.58 (0.53 – 0.63) | 0.74 (0.68 – 0.80) |
| Female | 0.56 (0.51 – 0.62) | 0.75 (0.69 – 0.82) |
| Age |  |  |
| <18 years | 1.02 (0.99 – 1.05) | 0.93 (0.83 – 1.04) |
| ≥18 years | 0.99 (0.98 – 1.01) | 0.94 (0.90 – 0.99) |
| Population density, persons per square Km |  |  |
| High >15000 | 0.54 (0.48 – 0.61) | 0.70 (0.63 – 0.78) |
| Low average <15000 | 0.59 (0.53 – 0.65) | 0.82 (0.75 – 0.90) |
| Proportion residing in slum areas |  |  |
| >50% | 0.58 (0.46 – 0.74) | 0.79 (0.63 – 0.98) |
| <50% | 0.57 (0.52 – 0.62) | 0.78 (0.72 – 0.84) |

<sup>a</sup> Pre-Lockdown (Feb 1 to March 24, 2020); Nationwide Lockdown (March 25 to May 31, 2020); Unlock 1 – 2 (June 1 to July 13, 2020); Regional Lockdown (July 14 to July 23, 2020); Unlock 2 (July 24 to July 31, 2020); Unlock 3 –4 (September 1 to September 30, 2020).
<sup>b</sup> Compared to Unlock 3 – 4.

### Health system preparedness

By the middle of the nationwide lockdown, 74 flu clinics dedicated to COVID-19 care were established in Pune. In addition, 6 COVID care centers and 3183 hospital beds were available for patients with asymptomatic/mild and severe COVID-19 disease, respectively.

### Geospatial distribution of the cumulative COVID-19 burden

Among the 15 ward offices shown in **Figure 1B**, the cumulative COVID-19 caseload steadily increased between pre-lockdown and post-lockdown phases (**Supplementary Video 1**). As illustrated in **Figure 1C**, initial spread was primarily confined to a few ward offices with comparatively higher population density. By the end of the nationwide lockdown, western Pune continued to report a low case burden compared to central and eastern parts of the city. By the end of the study period, the caseload in all ward offices was above 2000, but the largest burden remained in the central part of the city aligned along the north-south axis.

## Discussion

During the early stages of the COVID-19 pandemic, India implemented a historic nationwide 68-day lockdown among 1.3 billion people and Pune region implemented a 10-day regional lockdown for its 3.1 million people following a brief period of unlock to curb a rapid increase in new COVID-19 cases.[13, 25] By the week of September 1^st^ 2020, India ranked second in the cumulative number of COVID-19 cases globally[1], and Pune city had the country’s largest burden of active COVID-19 cases, overtaking other tier 1 cities.[26] This analysis of Pune city’s public health COVID-19 surveillance data found that the national lockdowns contributed to significant delay of the growth of cases by approximately 8 weeks. Furthermore, lockdowns flattened the COVID-19 pandemic curve with significant reductions in new cases and comparatively low infection rates during the nationwide and regional lockdown. Rapid expansion of cases was observed during later unlock periods corresponding to resumption of normal economic activities, following a pattern similar to countries that did not impose mobility restrictions[1]. In addition, we found that the health system capacity was rapidly optimized during complete lockdown with flu clinic, COVID-care center and hospital bed expansion. Overall, this report provides real-world evidence of the impact of lockdowns on both the spread of a highly transmissible infectious disease and the strengthening of health systems in an urban area of India with variable population density.

The flattening of COVID-19 epidemic curves during nationwide and regional lockdowns was uniform across age groups and population densities. The greatest impact of the nationwide lockdown was observed among ward offices with high population density characterized by a 2% weekly decrease in new cases. The Pune regional lockdown appeared to be most effective among ward offices with >50% of the population residing in slum areas, as evidenced by a 13% weekly decrease in new cases. Furthermore, early post lockdown period witnessed slower rise in weekly incident COVID-19 cases indicating the continued impact of lockdowns on maintaining social distancing. These findings are consistent with early modelling studies based on Indian nationwide data up to mid-April, which predict the effectiveness of strict social distancing for 42 days to reduce incident COVID-19 cases.[13] Population-wide data from Spain and Italy also demonstrate that strict lockdown was successful in flattening the COVID-19 epidemic curve.[27] However, this evidence may not be applicable to India and other resource-limited settings, as European lockdown periods were shorter at 14 – 21 days, and population densities and dynamics are significantly different.[28]

Consistent with earlier reports from India and other resource-rich settings, our analysis indicates that advancing age increases the risk of both contracting COVID-19 disease and mortality.^3-7^ However, we did not find a sex-specific predilection for SARS COV-2 infection. We found that children <5 years had the lowest rate of infection, yet our observed infection rate appears to be much higher than limited prior reports.[1, 3-7] Notably, our geospatial visualization of the cumulative COVID-19 burden confirms the relatively rapid community-wide spread of the infection during the unlock periods.[7] The average population density in Pune City, 5600 persons per square kilometer, is more than 10-fold higher than the average population density in India (464 persons per square kilometer.[28, 29] Thus, it is not surprising that the geospatial distribution of the COVID-19 burden was staggeringly rapid in Pune, particularly in the highest population density ward offices, which has contributed to India’s growing case counts.

The main strength of our study is the generalizability of the evidence to other more than 60 tier 2 cities in India and worldwide that have similar but varying ranges of population density within urban regions. Because the PMC surveillance data included individual patient addresses, we were able to assess the impact of lockdown in areas with differing population density within the city as well as map the geospatial distribution of COVID-19 cases before, during and after lockdown periods. However, this analysis included 44% of the original dataset that had after excluding records with missing data.[7] Although nasopharyngeal swab PCR was the most common SARS CoV-2 diagnostic used in the public health system, around 35% of tests were done using rapid antigen testing. As the rapid antigen test is known to have high specificity and low sensitivity[30], the case burden may have been underestimated when this testing method was used. Finally, risk of infection and mortality of COVID-19 among comorbid groups (diabetes, hypertension) could not be assessed, as these data were not captured.

In conclusion, as randomized controlled trial is not possible, this analysis of COVID-19 public health surveillance data is very relevant with re-emergence of a spike of cases in India which likely represents a second wave and/ or emergence of SARS-CoV-2 variants [31]. Importantly, this real-world data provides robust evidence that strict lockdowns to ensure physical distancing can be an effective public health measure to reduce the spread of highly transmissible infectious diseases, such as SARS-CoV-2 infection. In addition, the early extended lockdown in India may have facilitated the strengthening of health systems – a critical component of the public health response to any pandemic – as demonstrated by increased testing and hospital/critical care beds over time in Pune. With highly transmissible mutant strain of SARS-CoV-2 first detected in England, South Africa but rapidly spreading across the world[32, 33], our study adds confidence that non-pharmacologic interventions can be re-instated to limit its transmission in resource-limited settings. Further studies are needed to assess the epidemiology and effectiveness of lockdown in semi-rural and rural regions of India and beyond to inform successful, universal strategies for resource-limited settings worldwide.

## Data Availability

The data will be available upon a relevant request.

## Acknowledgements

We thank the Pune Knowledge Cluster principal investigators, Drs. Shashidhara and Ajit Kembavi, for their support in data acquisition. We thank Drs. Jonathan Golub, Akshay Gupte, Amita Gupta, Brial Wahl and Shashidhara for critical review of the paper and Dr. Katherine McIntire for copy-editing. We acknowledge generous support and congratulate PMC commissioner, officials and staff involved in the successful COVID-19 surveillance program. BSP would like to thank Dr. Snehal Shekatkar for discussion.

## Author contributions

VM, JM, NG, PB conceived the study. Data management and curation was performed by AS, JM. Geospatial mapping was created by JM. BSP conducted the modelling analysis; NG conducted the data analysis and all authors contributed to data interpretation and manuscript writing.

## Supplementary Material

**Supplementary Table S1.** Official name and number corresponding to each ward office located within Pune Municipal Corporation.

| Ward Number | Ward Name |
| --- | --- |
| 1 | Nagar Road Vadgaonsheri |
| 2 | Yerwada-Kalas-Dhanori |
| 3 | Dhole Patil Road |
| 4 | Aundh-Baner |
| 5 | Shivajinagar-Ghole Road |
| 6 | Kothrud-Bawdhan |
| 7 | Dhankawadi-Sahakarnagar |
| 8 | Sinhagad Road |
| 9 | Warje-Karvenagar* |
| 10 | Hadapsar-Mundhawa |
| 11 | Wanawadi-Ramtekdi |
| 12 | Kondhwa-Yewalewadi |
| 13 | Kasba-Vishrambagwada* |
| 14 | Bhawani Peth* |
| 15 | Bibwewadi* |

\*Denotes highest quartile of population density

**Supplementary Material Video 1. Video clip of geospatial mapping of daily COVID-19 caseloads over the analysis period**.

**Supplementary Material Methods. Geocoding methodology**

## References

1. Dong E DH, Gardner L. : An interactive web-based dashboard to track COVID-19 in real time. The Lancet Infectious Diseases. 2020;20(5):533–4.

2. Zhu N, Zhang D, Wang W, Li X, Yang B, Song J, Zhao X, Huang B, Shi W, Lu R et al: A Novel Coronavirus from Patients with Pneumonia in China, 2019. The New England journal of medicine 2020, 382(8):727–733.

3. Zhou F YT, Du R, Fan G, Liu Y, Liu Z, et al. :Clinical course and risk factors for mortality of adult inpatients with COVID-19 in Wuhan, China: a retrospective cohort study. The Lancet. 2020;395(10229):1054–62.

4. Richardson S HJ, Narasimhan M, Crawford JM, McGinn T, Davidson KW, et al. :Presenting Characteristics, Comorbidities, and Outcomes Among 5700 Patients Hospitalized With COVID-19 in the New York City Area. JAMA. 2020. May 26;323(20):2052–2059.

5. Clark A JM, Warren-Gash C, Guthrie B, Wang HHX, Mercer SW, et al. :Global, regional, and national estimates of the population at increased risk of severe COVID-19 due to underlying health conditions in 2020: a modelling study. Lancet Glob Health. 2020 Aug;8(8):e1003–e1017. doi: 10.1016/S2214-109X(20)30264-3. Epub 2020 Jun 15. PMID: 32553130; PMCID: PMC7295519.

6. Docherty AB HE, Green CA, Hardwick HE, Pius R, Norman L, et al. :Features of 20 133 UK patients in hospital with covid-19 using the ISARIC WHO Clinical Characterisation Protocol: prospective observational cohort study. BMJ. 2020 May 22;369:m1985. doi: 10.1136/bmj.m1985. PMID: 32444460; PMCID: PMC7243036.

7. Laxminarayan R WB, Dudala SR, Gopal K, Mohan B C, Neelima S, Jawahar Reddy KS, Radhakrishnan J, Lewnard JA. : Epidemiology and transmission dynamics of COVID-19 in two Indian states. Science. 2020 Nov 6;370(6517):691–697. doi: 10.1126/science.abd7672. Epub 2020 Sep 30. PMID: 33154136.

8. Kaur SP GV: COVID-19 Vaccine: A comprehensive status report. Virus Res. 2020 Oct 15;288:198114. doi: 10.1016/j.virusres.2020.198114. Epub 2020 Aug 13. PMID: 32800805; PMCID: PMC7423510.

9. Thanh Le T AZ, Kumar A, Gómez Román R, Tollefsen S, Saville M, Mayhew S. : The COVID-19 vaccine development landscape. Nat Rev Drug Discov. 2020 May;19(5):305–306. doi: 10.1038/d41573-020-00073-5. PMID: 32273591.

10. Dhama K SK, Tiwari R, Dadar M, Malik YS, Singh KP, Chaicumpa W. : COVID-19, an emerging coronavirus infection: advances and prospects in designing and developing vaccines, immunotherapeutics, and therapeutics. Hum Vaccin Immunother. 2020 Jun 2;16(6):1232–1238. doi: 10.1080/21645515.2020.1735227. Epub 2020 Mar 18. PMID: 32186952; PMCID: PMC7103671.

11. Agarwal A MA, Kumar G, Chatterjee P, Bhatnagar T, Malhotra P; PLACID Trial Collaborators. : Convalescent plasma in the management of moderate covid-19 in adults in India: open label phase II multicentre randomised controlled trial (PLACID Trial). BMJ. 2020 Oct 22;371:m3939. doi: 10.1136/bmj.m3939. Erratum in: BMJ. 2020 Nov 3;371:m4232. PMID: 33093056; PMCID: PMC7578662.

12. Rayner CR SP, Hershberger K, Wesche D. : Optimizing COVID-19 Candidate Therapeutics: Thinking Without Borders. Clin Transl Sci. 2020 Sep;13(5):830–834. doi: 10.1111/cts.12790. Epub 2020 May 2. PMID: 32212378; PMCID: PMC7485940.

13. Ray D SM, Bhattacharyya R, Wang L, Du J, Mohammed S, Purkayastha S, et al. :Predictions, role of interventions and effects of a historic national lockdown in India’s response to the COVID-19 pandemic: data science call to arms. Harv Data Sci Rev. 2020;2020(Suppl 1):10.1162/99608f92.60e08ed5. doi: 10.1162/99608f92.60e08ed5. Epub 2020 Jun 9. PMID: 32607504; PMCID: PMC7326342.

14. Giordano G BF, Bruno R, Colaneri P, Di Filippo A, Di Matteo A, Colaneri M. : Modelling the COVID-19 epidemic and implementation of population-wide interventions in Italy. Nat Med. 2020 Jun;26(6):855–860. doi: 10.1038/s41591-020-0883-7. Epub 2020 Apr 22. PMID: 32322102; PMCID: PMC7175834.

15. Chu DK AE, Duda S, Solo K, Yaacoub S, Schünemann HJ; COVID-19 Systematic Urgent Review Group Effort (SURGE) study authors. : Physical distancing, face masks, and eye protection to prevent person-to-person transmission of SARS-CoV-2 and COVID-19: a systematic review and meta-analysis. Lancet. 2020 Jun 27;395(10242):1973–1987. doi: 10.1016/S0140-6736(20)31142-9. Epub 2020 Jun 1. PMID: 32497510; PMCID: PMC7263814.

16. Gilbert M PG, Pinotti F, Valdano E, Poletto C, Boëlle PY, et al. :Preparedness and vulnerability of African countries against importations of COVID-19: a modelling study. Lancet. 2020 Mar 14;395(10227):871–877. doi: 10.1016/S0140-6736(20)30411-6. Epub 2020 Feb 20. PMID: 32087820; PMCID: PMC7159277.

17. Walker PGT WC, Watson OJ, Baguelin M, Winskill P, Hamlet A, Djafaara BA, et al.: The impact of COVID-19 and strategies for mitigation and suppression in low- and middle-income countries. Science. 2020 Jul 24;369(6502):413–422. doi: 10.1126/science.abc0035. Epub 2020 Jun 12. PMID: 32532802; PMCID: PMC7292504.

18. Lahiri A JS, Bhattacharya S, Ray S, Chakraborty A. : Effectiveness of preventive measures against COVID-19: A systematic review of In Silico modeling studies in indian context. Indian J Public Health. 2020 Jun;64(Supplement):S156–S167. doi: 10.4103/ijph.IJPH_464_20. PMID: 32496248.

19. Group. RC-S: Combating the COVID-19 pandemic in a resource-constrained setting: insights from initial response in India. BMJ Glob Health. 2020 Nov;5(11):e003416. doi: 10.1136/bmjgh-2020-003416. PMID: 33187963; PMCID: PMC7668115.

20. dashboard. PMCC-iP: http://cms.unipune.ac.in/~bspujari/Covid19/Pune2/. Accessed on 15 December 2020.

21. Hazra et. al: “INDSCI-SIM A state-level epidemiological model for India”, Private Communication [2021] Note: https://indscicov.in/indscisim.

22. B. S. Pujari, S Sheshatker: “Multi-city modeling of epidemics using spatial networks: Application to 2019-nCov (COVID-19) coronavirus in India”. medRxiv 2020.03.13.20035386 [2020]. https://doi.org/10.1101/2020.03.13.20035386.

23. Hilton J KM: Estimation of country-level basic reproductive ratios for novel Coronavirus (SARS-CoV-2/COVID-19) using synthetic contact matrices. PLOS Computational Biology 16(7): e1008031.

24. Sinha S: Epidemiological dynamics of the COVID-19 pandemic in India: an interim assessment. Stat. Appl, 18, 333–350.

25. Laxminarayan R WB, Dudala SR, Gopal K, Mohan B C, Neelima S, et al. :Epidemiology and transmission dynamics of COVID-19 in two Indian states. Science. 2020 Nov 6;370(6517):691–697. doi: 10.1126/science.abd7672. Epub 2020 Sep 30. PMID: 33154136.

26. Newspaper. HT: Pune district has highest Covid-19 case count in India. September 3, 2020.

27. A. T: Evaluation of the lockdowns for the SARS-CoV-2 epidemic in Italy and Spain after one month follow up. Sci Total Environ. 2020 Jul 10;725:138539. doi: 10.1016/j.scitotenv.2020.138539. Epub 2020 Apr 6. PMID: 32304973; PMCID: PMC7195141.

28. Worldometer: Countries in the world by population (2020). https://www.worldometers.info/world-population/population-by-country/.

29. Corporation. PM: https://www.pmc.gov.in/en/census. Accessed 11th December 2020.

30. Resource. C: Interim guidance for Antigen Testing for SARS-CoV-2. https://www.cdc.gov/coronavirus/2019-ncov/lab/resources/antigen-tests-guidelines.html.

31. Mint. N: India bracing up for the second wave of Covid-19 pandemic. February 22, 2021. https://www.livemint.com/news/india/india-bracing-up-for-the-second-wave-of-covid-19-pandemic-11613999656468.html.

32. E. M: Covid-19: What have we learnt about the new variant in the UK? BMJ. 2020;371:m4944. doi:10.1136/bmj.m4944.

33. Mustapha JO AI, Ajagbe OOR, Emeribe AU, Fasogbon SA, Onoja SO, et al. :Understanding the implications of SARS-CoV-2 re-infections on immune response milieu, laboratory tests and control measures against COVID-19. Heliyon. 2021 Jan 9;7(1):e05951. doi: 10.1016/j.heliyon.2021.e05951. PMID: 33490695; PMCID: PMC7810769.

